# A control framework to optimize public health policies in the course of the COVID-19 pandemic

**DOI:** 10.1101/2021.01.28.21250692

**Authors:** Igor M L Pataro, Juliane F Oliveira, Marcelo M Morato, Alan A S Amad, Pablo I P Ramos, Felipe A C Pereira, Mateus S Silva, Daniel C P Jorge, Roberto F S Andrade, Maurício L Barreto, Marcus Americano da Costa

## Abstract

The SARS-CoV-2 pandemic triggered substantial economic and social disruptions. Mitigation policies varied across countries based on resources, political conditions, and human behavior. In the absence of widespread vaccination able to induce herd immunity, strategies to coexist with the virus while minimizing risks of surges are paramount, which should work in parallel with reopening societies. To support these strategies, we present a predictive control system coupled with a nonlinear model able to optimize the level of policies to stop epidemic growth. We applied this system to study the unfolding of COVID-19 in Bahia, Brazil, also assessing the effects of varying population compliance. We show the importance of finely tuning the levels of enforced measures to achieve SARS-CoV-2 containment, with periodic interventions emerging as an optimal control strategy in the long-term.

**One-sentence summary:** We present an adaptive predictive control algorithm to provide optimal public health measures to slow the COVID-19 transmission rate.

## Introduction

Efforts to mitigate and circumscribe the spread of the SARS-CoV-2 virus have so far relied on non-pharmacological interventions (NPIs), set in place by most countries since March 2020, including social distancing, personal protective measures, mass quarantines, and other forms of limiting population movement. While the timely deployment of measures combined with an appropriate breadth of interventions has proven successful in effectively reducing the transmission rates of this virus (*1,2*), the socioeconomic impacts caused by extensive lock-downs are notably harsher to lower- and middle-income countries (*3, 4*), for which rescue spending packages and other bailout plans are unforeseeable. In this sense, finding ways to establish an equilibrium between keeping transmission under control while minimizing damages to the economy and society is highly desirable.

The challenges involved in controlling the SARS-CoV-2 epidemic are many. Using Brazil as an example, it is a very large country that ranks third in the number of reported COVID-19 cases (after the US and India), registering over 8.1 million infections and more than 203,000 deaths (*5*). First-wave mitigation strategies were largely decentralized, and the majority of governmental interventions occurred by local actions taken by the 26 states and the federal district, and their 5,570 municipalities (*6, 7*). Still, mitigation efforts were inadequate to keep SARS-CoV-2 transmission under control, and the collapse of health services was described throughout the country, which probably influenced the number of fatal outcomes observed to date (*8–12*). Even though in some areas of the country a very high level of infection was reached, such as the city of Manaus with an attack rate of 76% (*13*), this was insufficient to prevent new waves of infection, confirming that herd immunity is not a feasible or ethical route to tackle COVID-19 (*14, 15*). Thus, it is a concrete possibility that subsequent epidemic waves could, once again, pose a heavy burden on health services with consequent loss of lives, in line with the recrudescence of transmission observed in many countries, possibly boosted by the resuming of many economic and social activities.

Mathematical models have played a key role in assessing the effectiveness of public health policies and NPIs to contain the spread of SARS-CoV-2, as well as to evaluate the transmission dynamics of COVID-19 and how it is impacted by the movement of people (*2, 7, 16–20*). However, the bridging of model outputs to governmental actions aimed at reducing mobility is limited by the inherent uncertainties surrounding the obtained estimates, interpretation difficulties by policy-makers, and the lack of full understanding of a model’s predictive capabilities and limitations (*21*).

Accordingly, control algorithms coupled to epidemiological models provide an intuitive means to derive health policies and NPIs from data (*22–25*). By drawing on the availability of widespread mobility traces from cell phones, and building on the premise that circulation of individuals is a chief contributing factor for SARS-CoV-2 transmission (*26*), here we report an adaptive Nonlinear Model Predictive Control (NMPC) strategy able to reliably predict an optimal level of governmental interventions to decrease mobility, considering different degrees of social mobility effects, that reduces cases and fatalities and keeps hospitalization requirements below their limits while averting the unnecessary extension of restrictive measures such as lock-downs. We applied the NMPC algorithm to study the disease dynamics in Bahia, the largest and most populous state of Northeast Brazil, with territorial extension comparable to that of France. This framework, however, could be adaptable to deploy in multiple settings and can be particularly useful to other developing nations, which lack the purchasing power of high-income countries to benefit from early vaccine access (*27*), and thus will probably have to coexist with the pandemic effects for longer.

## Results

We present our results under the framework of nonlinear disease spread modeling coupled with control theory methods (*28*). This strategy is sufficiently general to be applied to different settings and can be replicated with minimal data information requirements, which are available for most other countries, ie. cases, fatalities, and hospitalization occupancy beds. To illustrate the utility of the method, we subdivide the following sections toward studying the transmission dynamics in the state of Bahia, Brazil. Three steps are described: 1) the definition of a compartmental model that expresses the dynamic of cases, fatalities, and health service requirements, taking into account asymptomatic/non-detected cases and social mobility patterns; 2) an extension of this compartmental model by a system identification procedure including optimal gains to improve forecast accuracy; 3) the inclusion of an optimal control algorithm that can reliably direct the lifting, continuation or intensification of NPIs in light of the epidemiological situation (Fig. S8). Throughout the text, we use the terms public health policies and NPIs interchangeably to refer to government measures aimed at controlling COVID-19. We were particularly interested in studying policies that resulted in changes to population mobility patterns, since the circulation of (possibly infected) individuals, including a-/oligo-symptomatics, is a key factor sustaining the spread of SARS-CoV-2. We refer to this subset of policies as social distancing measures.

### Enactment of governmental measures and implications on mobility patterns

Considering the large territory and population of Bahia (Fig. S1), the COVID-19 dynamics in this state is comparable to that of a whole country. On March 6, the first case was registered in the state, roughly one week after the first confirmed case in the country. By September 15, a total of 285,448 cases had been confirmed, of which 6,040 resulted in deaths. On March 16, the state government established a set of measures to mitigate the transmission. They were implemented and partially eased during the period, primarily targeting specific regions rather than the entire state. Most adopted interventions are related to the restrictions of public events and closure of schools/universities. We note that, more detailed information about each government measure is discussed by Jorge et al. (*7*) and described in table S1.

Initially, we assessed whether there existed a relationship between the extent of government policies and mobility patterns. For this, the stringency index (*u*), and the social mobility reduction index (SMRI) were used, respectively, as proxies to measure the “strength” of the public policies and the consequent degree of population compliance (Fig. 1). The SMRI had a baseline average of 28.5% (February 1-28). In what followed, six characteristic temporal states were identified: 1) March 6-15: community transmission had been declared in the state, but no governmental measure had been established (stringency *u* = 0; average SMRI *ψ* = 30.9%; 2) March 16-20: initial measures were set in place (*u* = 33.7%; average SMRI *ψ* = 34.4%); 3) March 21-May 10: this period corresponded to the peak population adherence to distancing recommendations (*u* = 40.9%; average SMRI *ψ* = 46.0%); 4) May 11-June 15 and June 25-August 20: *u* reached its maximum value of 49.2% (representing the peak of interventions) with average SMRI of 42.2% and 41.2%, respectively; 5) June 6-23: population adherence started to decrease with the concurrent reduction in stringency (*u* = 40.9%; average SMRI *ψ* = 39.1%); and 6) August 21-September 15: following the peak number of cases, a progressive decrease of both stringency and adherence ensued (*u* = 39.1%; average SMRI *ψ* = 39.0%).

**Fig. 1.**
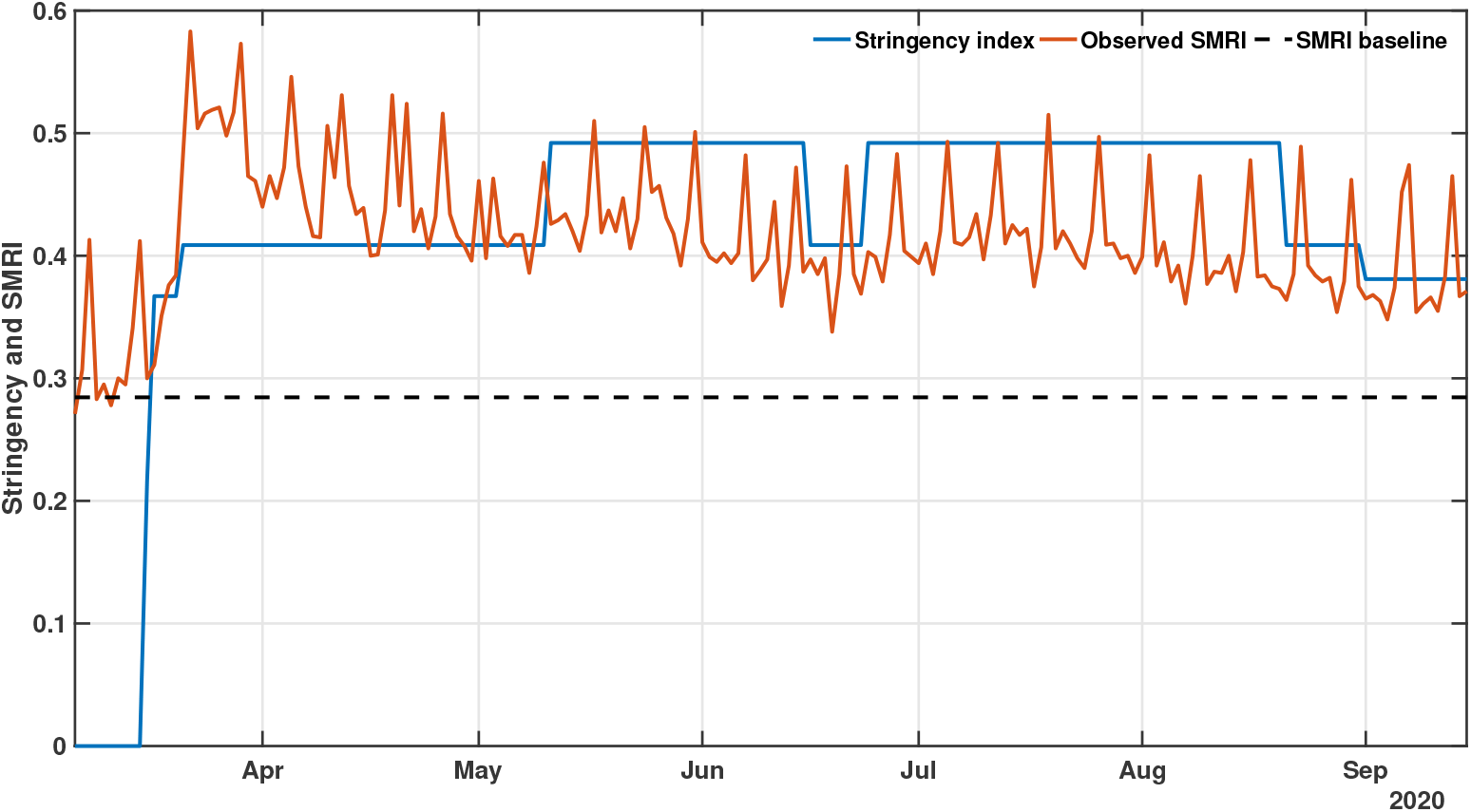
COVID-19 governmental polices and population response. The plot shows the stringency (blue) and social mobility reduction (orange) indexes in Bahia. Raw data from March 6 to September 15, 2020 are shown in this graph. The dashed horizontal line represents the baseline SMRI average between February 1-28, 2020.

### Reproducing the dynamics of COVID-19

To reproduce the transmission dynamics of COVID-19 in Bahia, under the previous described social behavior and governmental interventions, we applied the SEIIHURD+*ψ* model with all gains *g*_*i*_ = 1 in supplementary equations (1a) to (1h). A sensitivity analysis of the model was performed, allowing identification of key parameters governing the dynamics of this system (detailed in Supplementary Text).

Based on a visually good fit between observed and model-predicted values, the SEIIHURD+*ψ* model can reproduce the dynamics of COVID-19 with respect to the number of cases, deaths, and clinical hospitalization/ICU bed requirements (Fig. 2). In this simulation, epidemiological parameters of the model were kept fixed (table S2), while the SMRI (given by the registered series *ψ*) and the transmission rate *β* varied in time. The goodness of fit, estimated using *R*^2^, varied between 0.4684 (for the number of cases) and 0.9844 (for the ICU requirements). We next aimed to improve the forecast accuracy of the model, in particular for the prediction of cases, by employing a parameter identification procedure.

**Fig. 2.**
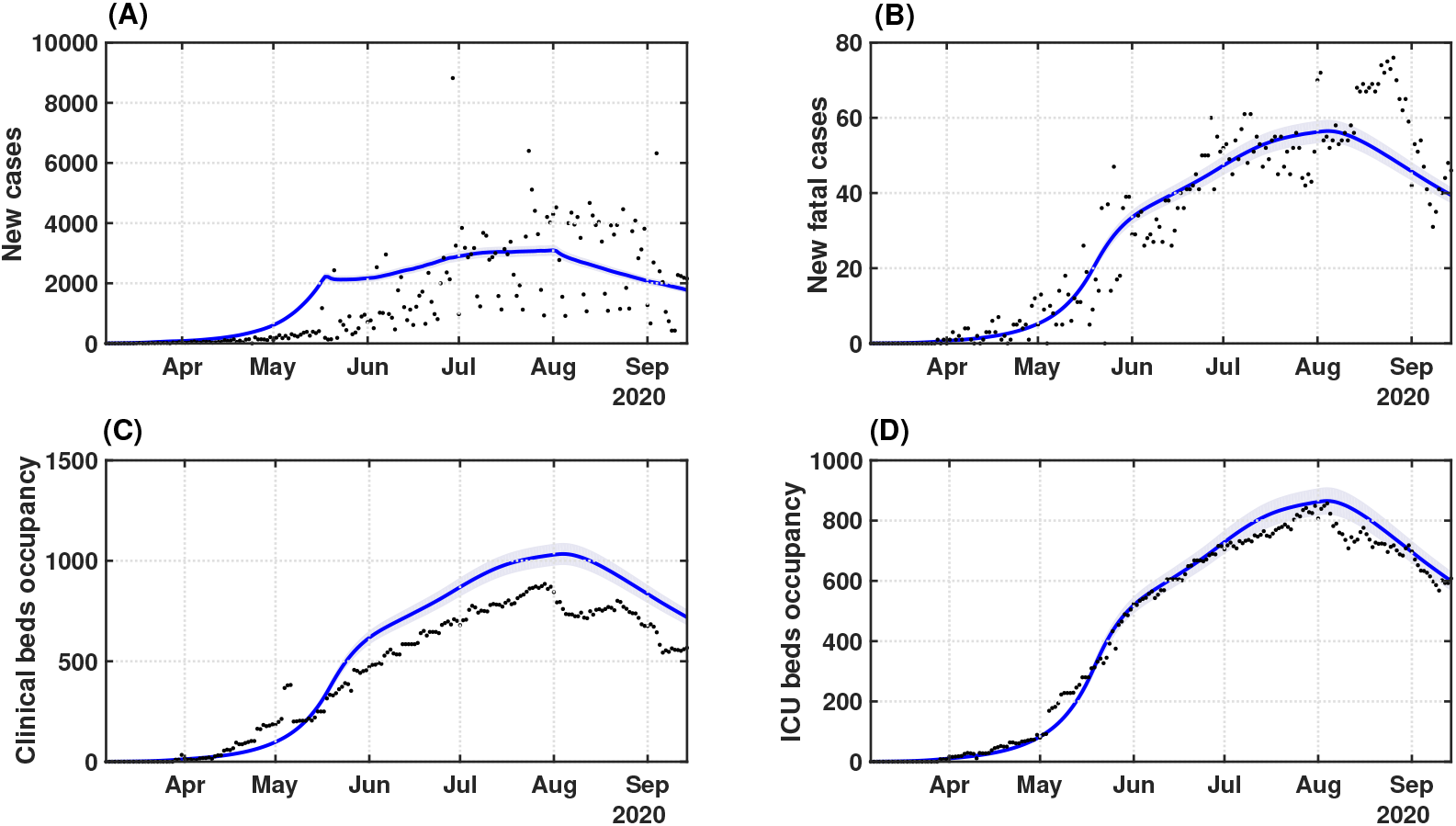
Transmission dynamics of COVID-19 in Bahia. Effects of the implemented interventions, mobility patterns and respective coefficient of determination *R*^2^ on the dynamic of (A) cases (*R*^2^ = 0.4684), (B) deaths (*R*^2^ = 0.8871), (C) clinical hospitalization (*R*^2^ = 0.7953) and (D) ICU bed requirements (*R*^2^ = 0.9844) at the state level. The solid blue lines represent the evolution of the epidemic given by the SEIIHURD+*ψ* model without gains. The shaded error bands represent 5% of the curve extrapolation margin. The assumed parameter values are shown in table S2. Raw data (black dots) from March 6 to September 15, 2020 are shown in this graph.

### Improving forecasting of cases, deaths, and clinical/ICU occupancy

By drawing on the previous result, considering that the SEIIHURD+*ψ* model with unitary gains can realistically describe the COVID-19 transmission dynamics, we sought to couple an internal controller capable of predicting optimal social mobility actions. Based on the control theory framework (*28*), the internal process in the NMPC algorithm requires an accurate forecast of the pandemic dynamics to formulate predictive control strategies based on a well-posed optimization problem.

Seeking to improve the forecast accuracy of the SEIIHURD+*ψ* model without compromising the epidemiological parameters, the gains *g*_*i*_ vary every 13 days, which is consistent with the infection dynamics of the SARS-CoV-2 virus (and could provide enough days for the validation tests). As an exception, the transmission parameter *β* may change in time, that is, the gain *g*_1_ is allowed to change within the 13 days windows. In particular, when the internal model also has *g*_*i*_ = 1, the results refer to a nominal case analysis. The main goal in the identification stage is to adapt the model fit by assuming that the input data series may suffer interference from several factors, such as case under-ascertainment and underreporting, as well as notification delays (*29*). In addition, the model parameters may undergo slight variations in the unfolding pandemic due to changes in medical treatments and protocols and the enactment of governmental measures, for instance. Consequently, we conducted an optimization stage to adjust the parameters of the SEIIHURD+*ψ* model and increase the quality of the predictions, particularly in the short-term, for which an enhanced accuracy benefits the most the results of the control algorithm.

Figure 3 presents the model validation, adjusted with data up to August 22, 2020, and its forecasts compared with real data for cumulative cases, deaths, clinical and ICU beds occupancy, up to September 15. The results from the identification approach show that, with a proper adjustment of the gains, it is possible to improve the model’s accuracy and offer reliable predictions for up to 25 days in the future. By assessing the coefficient of determination, *R*^2^, our results reveal that the optimized model can reproduce the series of cumulative cases (*R*^2^ = 0.9998), fatalities (*R*^2^ = 0.9994), clinical (*R*^2^ = 0.9872) and ICU bed requirements (*R*^2^ = 0.9872) with high accuracy (Fig. 3).

**Fig. 3.**
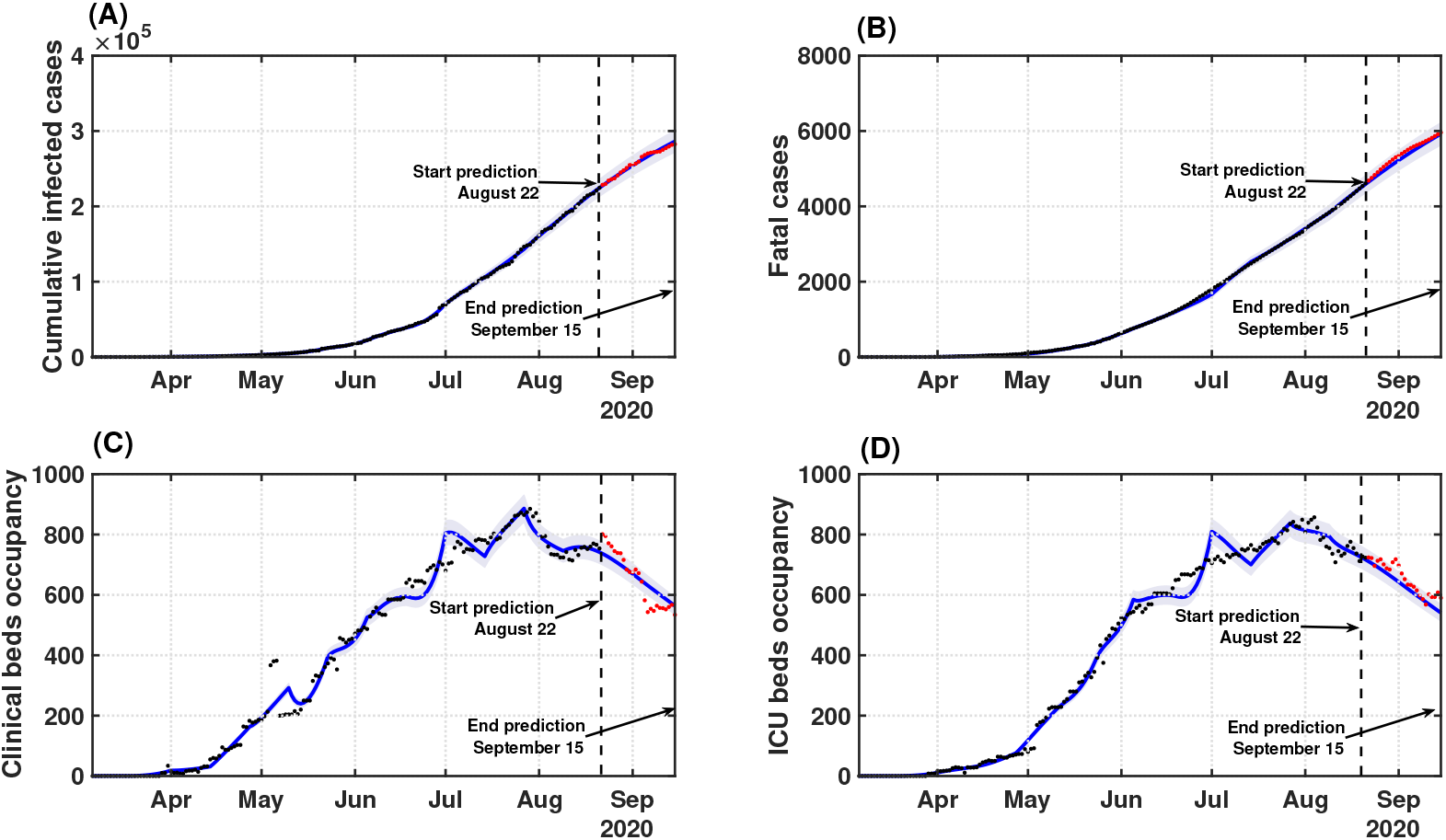
Model validation and forecast of the COVID-19 dynamics in Bahia. Model curves adjusted up to August 22 (blue lines), accounting for the identification procedure for *g*_*i*_ parameters, and respective coefficient of determination *R*^2^ for: (A) cumulative cases (*R*^2^ = 0.9998); (B) deaths (*R*^2^ = 0.9994); (C) clinical hospitalization (*R*^2^ = 0.9872) and (D) ICU bed requirements (*R*^2^ = 0.9872) at the state level. Data beyond the dashed vertical line indicate the predicted values for the epidemiological parameters between August 22 and September 15. The shaded error bands represent 5% of the curve extrapolation margin. The assumed model parameter values are shown in table S5. Raw data (black and red dots) from March 6 to September 15, 2020 are shown in this graph.

The estimated gains are shown in Tables S5 and S6. Although we obtained estimates for a total of 13 windows, the variance of the gain series is very small and enough to improve the forecast, indicating that the mathematical model is representative of the underlying epidemic unfolding process. By coupling the prediction capability of the SEIIHURD+*ψ* model with a control algorithm, an optimal framework for the deployment of governmental interventions that accounts for human mobility patterns can be developed.

### An optimal control guide for public health interventions

Next, we combined the SEIIHURD+*ψ* model with a predictive control algorithm in order to determine an optimal level of social mobility and governmental measures that would allow a reduction in cases and fatalities and preserve clinical and ICU bed occupancy rates below their limits. We note that the operation of this control algorithm is periodic, in such a way that new public health measures are determined every week, in line with the accurate short-term projections produced by the model. Also, many local governments now rely on multi-phased approaches to ease or increase the level of measures, usually based on objective metrics such as occupation of hospitals, *R*_*t*_, and trajectory dynamics of cases (*30–34*). Consequently, our strategy can also support the periodic re-calibration of stringency in phased reopening strategies to minimize the chances of surges.

Our control algorithm takes as input the time series of *u* and *ψ*, as presented in Fig. 1. The possible scenarios of people’s response influence the future values of stringency. As shown earlier, governmental measures impact population mobility, including in situations of low population compliance. This scenario is represented in Fig. 1, from early May to the end of August, when a downward trend in the SMRI over time is noted, despite increasing levels of *u*(*t*). Thus, we simulated three scenarios corresponding to high and low degrees of population compliance, translated into high/low mobility patterns, and a third scenario mimicking most closely what actually occurred during the period in terms of population mobility, therefore predicting the required levels of measures to reach epidemic control. For this latter scenario, the series of *ψ*(*t*) from March 6 to September 15 was given as input to the model. Since data for the SMRI from September 16 onward was unavailable, as we were projecting into the future, we considered that every three weeks the last SMRI value in the *ψ* time series would be lessened by 2% until reaching the minimum value measured at the beginning of the pandemic, reasoning that the decrease in the number of cases and deaths would lead to a reduction of the SMRI. We refer to this scenario as “validated model” in Fig. 5, since it uses the actual series of SMRI (truncated on September 16) shown in Fig. 4B.

**Fig. 4.**
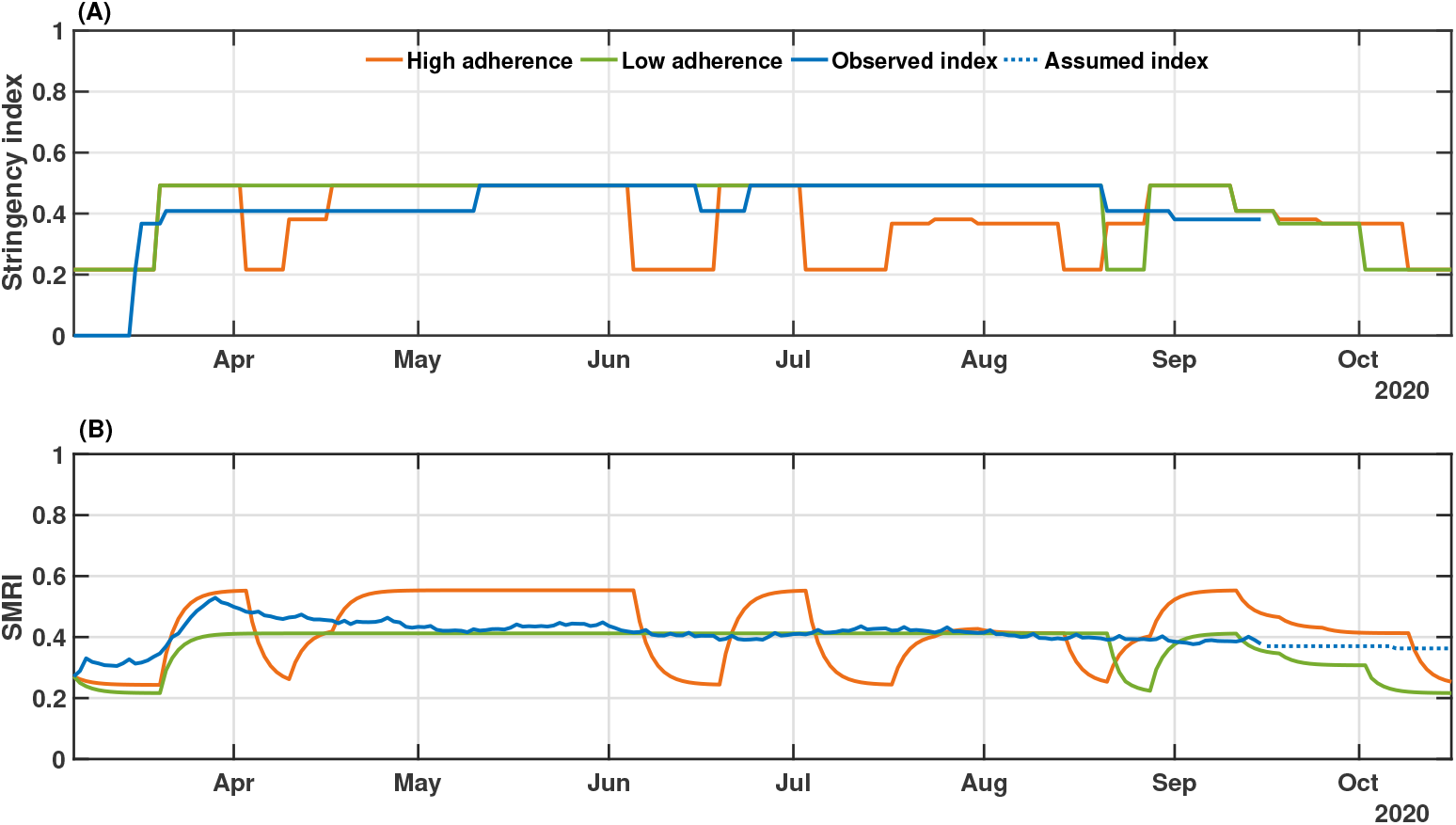
Real and simulated social mobility and governmental interventions in the state of Bahia. Levels of stringency (A) and social mobility reduction (B) indexes are shown for the high and low compliance scenarios, as well as the actual value of these metrics in the state-level during the period. The observed SMRI values (March 6-September 15) consist of a 8-day moving average. The dotted line in panel (B) indicates the assumed values of SMRI as described in Results.

**Fig. 5.**
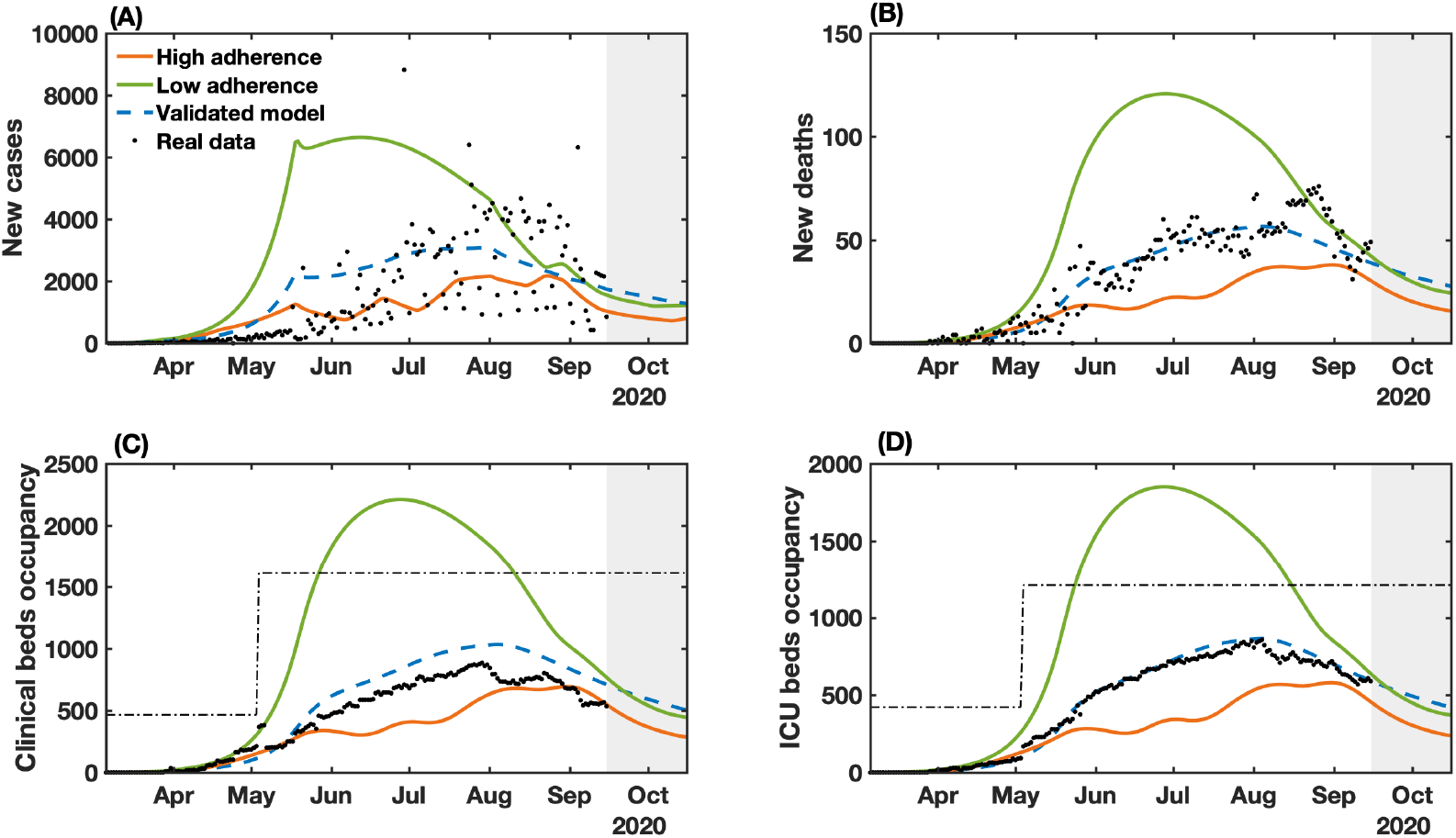
NMPC-controlled simulated epidemic unfolding compared to real-world data in Bahia, Brazil. (A) New cases; (B) deaths; (C) clinical hospitalization and (D) ICU bed requirements at the state level. The dashed-blue lines represent the dynamics of the validated model presented in Fig. 2 considering the observed SMRI time series in Fig. 3B. The dashed-dotted lines represent the clinical and ICU bed limits. Raw data (black dots) from March 6 to September 15, 2020 are shown in this graph.

High and low compliance scenarios were considered by adjusting supplementary equation (11), which provides the mathematical relation between *u* and *ψ*, evaluated on the historical data presented in Fig. 1. We calculated past gains *K*_*ψ*_ by considering the mean value of *ψ* in each constant period of *u*. Further, we considered only the data series beginning on March 16, when the first interventions were enacted. The calculated gains *K*_*ψ*_ are 1.0208 from March 16-20; 1.1247 from March 21-May 10; 0.8577 from May 11-June 15; 0.9560 from June 6-23; 0.8374 from June 24-August 20; and 0.9974 from August 21-September 15. Therefore, to simulate the control algorithm, we can define the high compliance scenario at *K*_*ψ*_ = 1.1247, corresponding to the dynamic at the beginning of the epidemic in the state; and the low compliance scenario at *K*_*ψ*_ = 0.8374, which corresponds to the period in which the peak of cases, fatalities, and hospitalizations starts to decline.

Another important factor to weigh is how stringent governments are willing to be in imposing control policies. While some countries opted for more liberal approaches to tackle COVID-19, such as Sweden, which relied mostly on responsible behavior, others maintained very strict curfews for extended periods (eg. Argentina) (*35*). A reasonable goal should be to keep the health care system below its capacity level, albeit our algorithm permits tuning the parameter *Q* to make stringency (*u*(*t*)) more or less flexible, which ultimately impacts economic sectors and social behavior. Therefore, from March 6 to May 15 we assume *Q* = 8 *·* 10^4^, from May 16 to August 23 we set *Q* = 3 *·* 10^4^ and from August 24 to October 15 *Q* = 1 *·* 10^4^. An analysis of this parameter’s variation and its effect on future predictions is given in Figs. S5 and S6.

In Fig. 4 we show the results of the NMPC algorithm applied to (re)construct the levels of governmental measures, that varied between 21.62% and 49.21%, enforced from March 6 to September 15, and a predicted scenario from September 16 to October 15, considering the popular adherence settings defined previously. The low and high compliance scenarios are compared with the levels of enacted measures since the start of the pandemic (Fig. 4A).

When we combine low popular compliance with the limited variation of the stringency index between 21.62% and 49.21%, the governmental measures must be maintained most of the time at their peak rate, only allowing for a relaxation at the end of August and up to September 3 until October 15, when the requirements of governmental measures reach their lowest degrees, according to the control algorithm. From the observed data, a stringency value of 49.21% corresponds to a two-third restriction on public events, the closure of all schools and universities, a one-quarter of government employees working from home, a 50% isolation compliance in areas subjected to lockdown, a closure of 28.6% non-essential activities and a one-quarter reduction in transportation (table S4). The low compliance scenario, presented in Fig. 4B, shows that a small variation on the mobility compared to the real-world data (black dashed line) could cause drastic changes on the epidemic curves shown in Fig. 5, almost doubling the number of infections and fatalities (by a factor of 1.84 and 1.89, respectively). As a result, full collapse of the health care system would occur earlier on the pandemic and the efforts made to expand the number of hospital beds in the state (represented by the increased bed capacity made available from May on; Fig. 4C and D) would not be sufficient for the requirements of COVID-19 patients.

In contrast, a more optimistic scenario is obtained with a high population compliance to measures. In Fig. 4A, we observe that when the population has a high level of adherence to measures, the optimal level of stringency can be kept under 40% for most of the time, with the highest values of stringency (49.21%) occurring between April 10 to May 21. To compare with the enforced measures, a 38.10% level of stringency index corresponds to the halting of 66.7% of public events, the closure of all schools and universities, one-quarter of government employees working from home and the closure of 42.8% of non-essential activities (*7*). In the high compliance scenario, the control framework yields an improved epidemiological situation compared with the actual scenario. The projection results in an estimated decrease of 38.2% in the number of cases, 37.9% in the number of deaths, and 33% of the maximum occupancy of clinical and ICU beds (Fig. 5). Our results also point that, in highly compliant populations, periodic interventions, i.e. alternating periods of high and low stringency, emerge naturally as the optimal strategy to promote control of the transmission (Fig. 4A). This can be a less dramatic mitigation alternative, which by itself could result in increased population adherence to measures.

The control results from September 16 to October 15 predicts an initial high level of required stringency in both low and high compliance scenarios (Fig. 4), followed by a gradual lifting while keeping the downward trend for the epidemic curves. This initial high-level requirement is intuitive if the aim is to reduce cases as close to the basal level as possible (dashed line in Fig. 5). In fact, in the low compliance scenario the number of cases, fatalities and hospitalizations is higher than the basal level represented, while in the high compliance scenario the transmission was still presenting a slow increase. The proposed control algorithm is able to provide the maximum allowed values of interventions to reduce the number of cases. However, the behavioral response is insufficient to allow a control of the epidemic curves as shown in Fig. 5. An alternative would be to strengthen government measures, in the case of low popular support, as shown in figs. S3 and S4.

Finally, comparing the control framework results with the unfolded local scenario, we note that the proposed control algorithm is able to maintain the clinical and ICU bed occupancy below the thresholds of available beds, reduce the total number of infection and deaths, while keeping approximately the same level of applied measures.

## Discussion

In this work, we introduced a framework for optimizing the required levels of public health policies, that translate into variable social distancing effects, during an unfolding pandemic. This tool combines control theory, parameter identification, and nonlinear dynamic modeling to optimize the level of governmental measures according to human behavior, in terms of mobility patterns, during a pandemic. We extend and validate a compartmental mathematical model (*9*) that describes the dynamic of symptomatic and asymptomatic/non-detected cases, deaths, and health service requirements, considering the temporal influence of social mobility. By evaluating the effects of social distancing measures enacted locally, we provide a mathematical relationship between interventions and the degree of compliance of the population, measured by the reduction in people’s mobility. We embedded this model in an adaptive control algorithm that can help set policy targets such as the maintenance, heightening or lifting of NPIs (Fig. 4). The utility of this approach is illustrated by studying the dynamics of COVID-19 in Bahia, Brazil, which offered opportunities for an enhanced control of the epidemic (Fig. 5). However, the method is simple and versatile and can be deployed to the analysis of other infectious diseases in other populations, predicting the level of required measures with accuracy. A benefit of this approach is that the levels of predicted stringency can be fine-tuned to adjust to the fragilities of the targeted population and the government capabilities to implement a measure, which can depend on multiple factors including the availability of local resources and political stability.

We used Bahia, Brazil as a proxy of an area with a large population, limited health infrastructure, and stark socioeconomic inequalities. First, we performed a pre-assessment to reproduce the dynamics of COVID-19 in the state from March 6 to September 15, 2020. By leveraging the real-world levels of locally implemented measures with the observed social mobility patterns in this period, our control algorithm identified optimal time windows where the measures and high level of population response (such as that observed at the beginning of the epidemic in the state) can be applied with different magnitudes. In this scenario, the number of cases, deaths and hospitalizations could have been averted by nearly 64%, and the population would have benefited with more extended periods of relaxation of social distancing measures through the enforcement of periodic interventions, which others have shown lead to improved transmission control (*36–38*), while alleviating the multiple deleterious facets of prolonged human confinements. Such an achievement can be strategically combined to reduce the impact on the health system and on the economy. In contrast, our findings highlight the importance of widespread adherence to enforced measures. In a scenario of low popular compliance, governments are forced to increase the level of measures to protect the health care system from collapsing. The results in Bahia show that a low compliance could lead to double the number of cases and deaths, and the accompanying collapse of the healthcare system would be inevitable. This is particularly important when planning measures in less developed countries, where poverty is associated with low education levels and, consequently, difficulties in realizing the importance of actions aimed at controlling spread of the virus (*39, 40*). More vigorous levels of stringency could further decrease the transmission rates; however, the economic effects of prolonged curfews cannot be ignored.

In practice, our proposal requires some care. In particular, long-term forecasting using mathematical models suffer from inherent uncertainties. A real-world application of our method would require constant re-calibration with newly observed data, which we showed substantially improved the accuracy of the predictions. Also, the underlying implemented model does not account for age structure, heterogeneity in contact patterns or stochasticity. In spite of its simplicity, the short-term predictions are still robust and thus adequate for supporting policy re-calibration in short time windows. Additionally, although we can predict the levels of stringency that should be applied in a region, more studies are warranted to understand how the different categories of intervention, such as the closure of schools, the limits on travels and people’s movements, among other NPIs, influence the reduction of cases, as well as how closely related interventions should be prioritized.

There has been strong interest in trying to define the set of NPIs most effectively capable of delaying the spread of COVID-19 (*1, 2, 16, 19, 41*). As noted, various factors complicate the choice of a particular measure over another: First, there is extensive overlap among commonly enacted NPIs (eg. ban on small- and large-scale gatherings); second, many measures were enacted in parallel or almost successively, hindering the evaluation of their individual effectiveness since they are highly correlated. Haug et al. (*2*) performed the most complete analysis of NPI effects to date, systematically measuring the impact on the *R*_*t*_ of COVID-19 of 6,068 individual measures in 79 countries and finding that no single intervention is able to reduce *R*_*t*_ below one, and that measures should be combined and deployed in a timely fashion for maximal efficacy. To avoid the pitfalls associated with having to determine the set of NPIs able to maximally decrease the growth of the epidemic, we instead focused on predicting an optimal level of stringency, that in turn influences the mobility patterns of the population. By plugging this relationship into a control algorithm, we were able to reliably assess the level of measures needed to reach a situation of epidemic control, particularly by averting full occupation of available hospital beds. It would be up to policy-makers to choose a combination of NPIs leading to the required stringency level, as predicted by the controller, since distinct combination of measures can lead to equivalent stringency values (*2*). More work is needed to better understand the value of individual NPIs and their optimal use to accomplish control targets.

Deploying this strategy in other settings would require, in addition to the local epidemiological data used during the calibration of part of the model’s parameters (*28*), more details of the population’s compliance to measures, as our results point that the level of adherence can markedly influence the dynamics of COVID-19 spread, and these may vary by factors such as education level and degree of individual freedom across countries. Consequently, identical levels of stringency may evoke different behavioral responses according to the compliance of each individual and their emerging collective attitudes–ie. people’s actions, including health behavior, are subject to multiple psychological factors and motivations (*42, 43*), herein modeled as high and low compliant populations. However, the compartmental model that serves as the basis for the dynamics of contagion is general enough to be readily used in other regions, while also being adaptable to other disease domains.

## Data Availability

The data that support the findings of this study are openly available in the repository of Center for Data and Knowledge Integration for Health (CIDACS) at https://github.com/cidacslab/covid-control-policies.

## Acknowledgments

We gratefully acknowledge the InLoco team, namely Jose Luciano Melo, Raiza Oliveira, Afonso Delgado, Abel Borges, Andre De’Carli, Hector Pinheiro, Lucas Rufino, Gabriel Teotonio and Luiza Botelho for providing raw files of the social mobility reduction data used here.

## Authors contributions

Conceptualization–JFO, IMLP, MAC, MLB, MMM, RFSA. Methodology–AASA, DCPJ, FACP, IMLP, JFO, MAC, MMM, MSS, RFSA. Writing - Original Draft—IMLP, JFO, MAC, RFSA. Writing - Review & Editing— IMLP, JFO, MAC, MLB, PIPR, RFSA.

## Data and materials availability

All data is available within the manuscript or its supplementary materials, as well as in the GitHub code repository at: https://github.com/cidacslab/covid-control-policies.

## Supplementary materials

Materials and Methods Supplementary Text Figs S1-S8

Tables S1-S5

References (44-49)

