## Supplementary Materials for "A control framework to optimize public health policies in the course of the COVID-19 pandemic"

January 26, 2021

**Abstract**

In this document we present the relevant Supplementary Materials accompanying the manuscript “A control framework to optimize social distancing measures in the course of the COVID-19 pandemic” by Pataro, Oliveira *et al.*

**This pdf file includes:**

Materials and Methods

Supplementary Text

Figs S1-S8

Tables S1-S5

### Materials and Methods

### Data collection

We collected epidemiological data of COVID-19 in Bahia, a state with 14.8 million population in the Northeast of Brazil, for the purpose of applying the model developed here. The data comprise the daily number of registered cases and deaths from COVID-19, as well as the daily number of clinical hospitalization and ICU requirements, obtained from the Secretary of Health of Bahia from March 6 to September 15, 2020. Social mobility is represented by the data from "InLoco", a Brazilian technology start-up, which is available at `http://mapabrasileirodacovid.inloco.com.br` (*44*). The mobility is measured according to the population behavior constructed from anonymous geo-movement information extracted from 60 million mobile devices throughout the country. The used metric, referred to in the manuscript as the social mobility reduction index (SMRI), is defined in the interval 0 to 100%. It measures the displacement of devices from its self-defined home location, such that the bigger the index, the lower is the population mobility.

In addition to the epidemiological and the social mobility data, we collected all the decrees aiming to mitigate the spread of the disease in the state of Bahia. This dataset resulted from an update to the effort originally described in work of Jorge at al. (*7*). Therein, the authors performed an analysis of all state governmental decrees applied in each Brazilian state, deriving an index to measure the stringency of COVID-19-associated interventions adapted to the Brazilian context, originally reported in (*45*). The index evaluate measures related to the Cancellation of public events (O1), Closure of schools/universities (O2), Home-office for governmental employees (O3), Isolation of groups or the whole population (O4), Closure of non-essential businesses and public activities (C1), and Transport lock (C2). As presented by Jorge at al. (*7*), the total stringency index is a combination of the evaluation of these measures. The index varies from 0 to 1 meaning that the lower the index, the lower the level of governmental measures to mitigate the spread of the disease in the region.

#### Model design

The SEIIHURD model considered here (*9*) describes the dynamics of a population divided into compartments as susceptible ($S$), exposed ($E$), asymptomatic/non-detected infections $(I_a)$ and symptomatic (reported) infections ($I_s$). The reported infections may present mild to severe symptoms, thus a proportion of them may require hospitalizations (clinical beds), ($H$), or intensive care unity (ICU) admission ($U$). After the infectious period, individuals may recover ($R$) from the disease. A more severe outcome results in death ($D$) due to COVID-19, after passing for a period of hospitalization or ICU. In this model the transmission is not affected by individuals in $H$ and $U$ compartments. Also, based on the results presented by Oliveira et al. (*9*), we also consider a flux of patients between the $H$ and $U$ compartments, modeling the condition that patients admitted to a clinical ward may worsen their condition and require an ICU bed; conversely, patients in intensive care may need clinical care prior to discharge and recovery.

We added new definitions to the SEIIHURD model to consider influences on the dynamics due to human behavior and healthcare improvements, by allowing the variation of some gains over time. The resulting model SEIIHURD+$\psi$ is described by the systems of equations (1) below:

$$\frac{dS}{dt} = \frac{-g_1(t)\beta(1-\psi(t))S(I_s + g_{11}(t)\delta I_a)}{N} \tag{1a}$$

$$\frac{dE}{dt} = \frac{g_1(t)\beta(1-\psi(t))S(I_s + g_{11}(t)\delta I_a)}{N} - \kappa E \tag{1b}$$

$$\frac{dI_a}{dt} = (1 - g_7(t)p)\kappa E - \gamma_a I_a \tag{1c}$$

$$\frac{dI_s}{dt} = g_7(t)p\kappa E - \gamma_s I_s \tag{1d}$$

$$\frac{dH}{dt} = g_2(t)hg_3(t)\xi\gamma_s I_s + (1 - g_4(t)\mu_U + g_8(t)\omega_U g_4(t)\mu_U)g_5(t)\gamma_U U - g_6(t)\gamma_H H \tag{1e}$$

$$\frac{dU}{dt} = g_2(t)h(1 - g_3(t)\xi)\gamma_s I_s + g_9(t)\omega_H g_6(t)\gamma_H H - g_5(t)\gamma_U U \tag{1f}$$

$$\frac{dR}{dt} = \gamma_a I_a + (1 - g_2(t)h\gamma_s I_s + (1 - g_{10}(t)\mu_H)(1 - g_9(t)\omega_H)g_6(t)\gamma_H H \tag{1g}$$

$$\frac{dD}{dt} = (1 - g_9(t)\omega_H)g_{10}(t)\mu_H g_6(t)\gamma_H H + (1 - g_8(t)\omega_U)g_4(t)\mu_U g_5(t)\gamma_U U \tag{1h}$$

Specifications of the epidemiological parameters are given in table S6. The temporal series $\psi$ accounting for social mobility patterns is given from the InLoco dataset previously described.

In order to improve the predictions that will dictate control measures, multiplicative gain factors, $g_i$'s, are introduced into some terms of the original SEIIHURD model. In this context, the factors $g_1, \ldots, g_{11}$ are used to account for temporal uncertainties in the parameters $\beta, h, \xi, \gamma_H, \gamma_U, \mu_H, \mu_U, \omega_H, \omega_U, \delta$ and $p$ according to the epidemic data which are read continuously by the control system. The uncertainties upon these parameters may appear over time due to different reasons during the course of pandemic, from noises and errors in the reported data to medical treatments improvements, equipment and medical supplies, and screening and testing measures. As a result, the internal adaptive control model can use these time-varying gains in order to enhance forecasting on the disease's spread and, thus, improve control performance (*22*,*24*). In contrast, when the terms $g_i$ are set to 1, the transmission rate $\beta$ is written in terms of a Heaviside step function as in Oliveira et al. (*9*), the equations [1a - 1h] reduce to the SEIIHURD model with social mobility index influencing the exposure to risks of the susceptible population, here defined as SEIIHURD+$\psi$ model with unitary gains.

#### Parameter estimation

To carry out our analysis, we consider the literature review presented by Oliveira et al. (*9*) to evaluate the key epidemiological parameters $\kappa$, $\gamma_a$ and $\gamma_s$, which are maintained fixed during the current work. The time-varying terms $g_1\beta$, $g_2h$, $g_3\xi$, $g_4\mu_U$, $g_5\gamma_U$, $g_6\gamma_H$, $g_7p$, $g_8\omega_U$, $g_9\omega_H$, $g_{10}\mu_H$ and $g_{11}\delta$ have the epidemiological search interval, shown in Table S3.

The estimation is performed by ordinary least square minimization problem in the identification algorithm so that the parameter values adjust the model to the series of cumulative cases ($C$), clinical beds occupancy ($H$), ICU beds occupancy ($U$) and deaths ($D$). The absolute error variables terms used in the optimization layer are the following:

$$Er_C(t) = |C(t) - \hat{C}(t, g_1(t), \ldots, g_{11}(t))|, \tag{2}$$

$$Er_H(t) = |H(t) - \hat{H}(t, g_1(t), \ldots, g_{11}(t))|, \tag{3}$$

$$Er_U(t) = |U(t) - \hat{U}(t, g_1(t), \ldots, g_{11}(t))|, \tag{4}$$

$$Er_D(t) = |D(t) - \hat{D}(t, g_1(t), \ldots, g_{11}(t))| \tag{5}$$

wherein the variables $\hat{C},\ \hat{H},\ \hat{U},\ \hat{D}$ are obtained according to the SEIIHURD+$\psi$ model equations [1a - 1h]. The complete optimization problem is formulated as follows:

$$\min_{g_i, \forall i=1,\ldots,11} \sum_{t=t_i}^{t=t_f} \left(w_1 Er_C(t) + w_2 Er_H(t) + w_3 Er_U(t) + w_4 Er_D(t)\right) \tag{6}$$

wherein $w_1 = 1,\ w_2 = 5$, and $w_3 = w_4 = 35$ are pre-selected positive weighting values (tuning parameters), used to normalize the order of magnitude of the total cost and minimize the errors $Er_C$, $Er_H$, $Er_U$ and $Er_D$. The values $t_i$ and $t_f$ define the interval $[t_i, t_f]$ where we want to apply the optimization layer.

When the SEIIHURD+$\psi$ with unitary gains is considered, the time-varying parameter $\beta$ is affected by the population mobility explicit in the model by the series of $\psi$. Thus, the values of

$\beta$ are estimated considering the mean value of $\psi$ over the identified interval of variations of $\beta$. Additionally, for the validation of the SEIIHURD+$\psi$ with unitary gains, we do not re-estimate the remaining parameters and they are kept as presented by Oliveira et al. (*9*) (table S2).

#### Model validation and prediction algorithm

In order to improve the performance of our future prediction, we separated our data into sets: a training set containing the first points, from $t_1$ to $t_{169}$, of each data series and a testing set containing the remaining points of the series going from $t_{170}$ to $t_{194}$, the last point of the data we are using. Within the training set, the data is split into consecutive two-week windows, which are sufficiently large to properly describe typical changes in social behavior. In each of these windows, we applied the optimization procedure defined in equation (6) to obtain gains $g_1\beta$, $g_2h$, $g_3\xi$, $g_4\mu_U$, $g_5\gamma_U$, $g_6\gamma_H$, $g_7p$, $g_8\omega_U$, $g_9\omega_H$, $g_{10}\mu_H$ and $g_{11}\delta$. We used the last estimated optimal values to forecast the remaining $25$ days of available data and demonstrate the validity of the forecast.

#### The control system

In the course of the COVID-19 pandemic, several measures were employed to overcome the socioeconomic impacts and the spread of SARS-CoV-2. The resulting control policies are passed either as a recommendation or as laws, aiming at reducing mobility, gatherings, and consequently slow the spread of the disease. Jorge et al. (*7*) evaluated the combination of restriction measures applied to each Brazilian state at each time $t$, yielding the stringency index of implemented governmental measures. The index varies from 0 to 1 depending on the level of restrictions, where $0$ means no COVID-19-specific measure applied and 1 corresponds to the most strengthened restriction, for instance, a full lockdown.

We denote here by $u(t)$ the value of the control signal (stringency index) at time $t$. The NMPC is formulated considering a prediction horizon $N_p$ of three weeks, i.e. $N_p = 21$ days. However, the optimal control signal is applied only on the first 7 days, being recalculated in the next week. We argue that it is not reasonable to change the measures in smaller periods, which would possibly cause confusion to the population.

Each future control signal $u(t)$ must be piece-wise constant and increase/decrease in accordance with plausible desired levels of control imposed on social mobility. Accordingly, we consider discrete values for $u$, as given in tables S4 and S5. Note that the control signal $u(t)$ lies in the interval $[0.2162\,,\,0.4921]$ for the scenario considering the real applied measures. For an hypothetical more rigorous scenario, the control signal lies in the interval $[0.2162\,,\,0.6269]$ (figs. 5 and S6). These values were obtained by matching the guidelines enacted in Bahia up to date, as provided by Jorge et al. (*7*). The algorithm to evaluate all possible future governmental measure is presented in NMPC algorithm section.

Bearing this in mind, we wanted to estimate what are the minimal measures necessary to guarantee that the number of clinical and ICU beds does not surpass the real capacity available in the state, that is, we seek to maintain:

$$H(t+j|t) \leq n_{\text{clinical}} \tag{7}$$

$$U(t+j|t) \leq n_{\text{ICU,}} \tag{8}$$

The information about the number of clinical and ICU bed available, denoted by $n_{clinical}$ and $n_{ICU}$, were obtained from the Secretary of Health of Bahia. The values were expanded in the course of the epidemic and it is given by $n_{\text{clinical}} = 466$ and $n_{\text{ICU}} = 422$ for the period between March 6 and May 4 (*9*), and $n_{\text{clinical}} = 1610$ and $n_{\text{ICU}} = 1210$ for the following period.

To maintain the desired levels of hospitalizations requirements given in Eqs. 7 and 8, the proposed NMPC algorithm must act in order to ensure the following control objective (trade-off objective):

$$J_{\text{NMPC}} = I_s(t_f) + Q \cdot u(t), \tag{9}$$

is obtained, wherein $Q$ is the weight tuning parameter used to ponder how much the control signal can vary. In other words, if $Q$ is low, $u(t)$ can vary more freely and achieve higher values, otherwise $u(t)$ achieves lower values. This trade-off adjustment is used to prioritize the minimization of either the control signal or the number of cases. Moreover, the tuning parameter $Q$ can also vary in time to adapt to the priorities and the epidemic situation. In this case, at the beginning, we restrict the control actions, aiming to simulate a more realistic scenario, in accordance to what was achieved by the initial enacted acts. Later, we start to relax the control action in order to reduce even more the number of infections. Lastly, when the number of infections is reducing, we reduce even more the value of $Q$, aiming to avoid subsequent waves of infection.

Of note, the control algorithm can be tuned to adjust to each situation, considering the stage of the pandemic, the level of occupancy of ICU and clinical beds and, mainly, the government's priorities, with a focus on increasing the restriction and the measures of social distancing or making them more flexible, adopting surveillance policies that work in parallel with the opening of economic activities and social life.

The control optimization process for the horizon of $N_p$ steps is assumed to minimize the number of symptomatic infection cases and also the stringency index measures, defined as follows:

$$\begin{aligned}
\min_{u(t)} J_{\text{NMPC}}(\cdot) &= \min_{u_t} \left( I_s(t_f) + Q \cdot u(t) \right), \qquad (10) \\
\text{s.t.} \quad & \text{SEIIHURD+}\psi \text{ Model } \forall\, i \in \mathbb{N}_{[1\,,\,N_p]}\,, \\
& \underline{u} \leq u(t+i-1) \leq \overline{u}\,, \\
& H(t+i) \leq n_{\text{beds}}\,, \\
& U(t+i) \leq n_{\text{ICU}}\,.
\end{aligned}$$

Given that the NMPC framework offers finitely parametrized social distancing guidelines for the COVID-19 spread, its implementation resides in simulating the validated SEIIHURD+$\psi$ along the prediction horizon with an explicit nonlinear solver and testing it according to all possible control $u$. Thus, the predicted variable $I_s$ at the last sample time $t_f$ of the prediction horizon $N_p$ is used to evaluate the cost function $J_{\text{NMPC}}(\cdot)$. The stringency index that implies in the violation of constraints are neglected. Then, the resulting control value is the one that yields the minimal $J_{\text{NMPC}}(\cdot)$, while abiding to the aforementioned constraints. Finally, the stringency index is applied and the horizon slides forward. This paradigm is explained in the NMPC Algorithm section. We note that this methodology ensures the optimality of the solution $u(k)$ regarding the control objective $J_{\text{NMPC}}$, as described by Rathai et. al. (*46*).

After calculating the optimal control signal, we consider that the population responds with a certain dynamic to governmental measures, as proposed by Morato et. al. (*25*). The dynamic response is defined as follows:

$$\psi(t+1) \;=\; \psi(t) + T_2 \varrho_\psi \left(K_\psi(t) u(t) - \psi(t)\right) \quad , \tag{11}$$

wherein $\varrho_\psi = 0.4\ \mathrm{day}^{-1}$ is a settling time parameter, which is related to the average time the population takes to respond to the enacted social isolation measures and $K_\psi$ is a gain relationship between $\psi$ and $u$. $T_2$ represents the sampling period of the NMPC algorithm of one week.

### Supplementary Text

### Parameter sensitivity analysis

We performed a sensitivity analysis to evaluate the effects of model parameters in the dynamics of $I_a$, $I_s$, $U$, $H$, $R$, $S$, $E$ and $D$ over time. By using an statistical variance-based method, described by Sobol (2001) (*47*), the sensitivity analysis of the system described by Eqs. (1a)-(1h) considers the following parameter vector

$$\theta := (\kappa, \gamma_a, \gamma_s, \gamma_h, \gamma_u, \mu_u, \xi, h, \mu_h, \omega_h, \omega_u, \delta, p) \in \mathbb{R}^{13}, \tag{12}$$

assuming that its elements are uniformly distributed in proper intervals:

$$\begin{array}{ccc} \kappa \sim \mathcal{U}(1/6, 1/3), & \gamma_a \sim \mathcal{U}(1/3.70, 1/3.24), & \gamma_s \sim \mathcal{U}(1/5, 1/3), \\ \gamma_h \sim \mathcal{U}(1/12, 1/4), & \gamma_u \sim \mathcal{U}(1/12, 1/3), & \mu_u \sim \mathcal{U}(0.4, 0.5), \\ \xi \sim \mathcal{U}(0.5, 0.99), & h \sim \mathcal{U}(0.05, 0.99), & \mu_h \sim \mathcal{U}(0.1, 0.2), \\ \omega_h \sim \mathcal{U}(0.1, 0.3), & \omega_u \sim \mathcal{U}(0.1, 0.3), & \delta \sim \mathcal{U}(0, 1.5), \\ & p \sim \mathcal{U}(0.13, 0.5). & \end{array} \tag{13}$$

To apply this method, we first generated sample values for the input factors shown in Eq. (12) by creating matrices $A$ and $B$, each with size $N \times n$, where $N$ is the number of samples and $n = 13$ is the number of parameters being analyzed, given by

$$A = \begin{pmatrix} \theta_1^{(A1)} & \theta_2^{(A1)} & \dots & \theta_i^{(A1)} & \dots & \theta_n^{(A1)} \\ \theta_1^{(A2)} & \theta_2^{(A2)} & \dots & \theta_i^{(A2)} & \dots & \theta_n^{(A2)} \\ \vdots & \vdots & \dots & \vdots & \dots & \vdots \\ \theta_1^{(AN)} & \theta_2^{(AN)} & \dots & \theta_i^{(AN)} & \dots & \theta_n^{(AN)} \end{pmatrix} \tag{14}$$

and

$$B = \begin{pmatrix} \theta_1^{(B1)} & \theta_2^{(B1)} & \dots & \theta_i^{(B1)} & \dots & \theta_n^{(B1)} \\ \theta_1^{(B2)} & \theta_2^{(B2)} & \dots & \theta_i^{(B2)} & \dots & \theta_n^{(B2)} \\ \vdots & \vdots & \dots & \vdots & \dots & \vdots \\ \theta_1^{(BN)} & \theta_2^{(BN)} & \dots & \theta_i^{(BN)} & \dots & \theta_n^{(BN)} \end{pmatrix}. \tag{15}$$

We then create $n$ matrices $A_B^i$, where column $i$ comes from matrix $B$ and all other $n-1$ columns come from matrix $A$:

$$A_B^i = \begin{pmatrix} \theta_1^{(A1)} & \theta_2^{(A1)} & \dots & \theta_i^{(B1)} & \dots & \theta_n^{(A1)} \\ \theta_1^{(A2)} & \theta_2^{(A2)} & \dots & \theta_i^{(B2)} & \dots & \theta_n^{(A2)} \\ \vdots & \vdots & \dots & \vdots & \dots & \vdots \\ \theta_1^{(AN)} & \theta_2^{(AN)} & \dots & \theta_i^{(BN)} & \dots & \theta_n^{(AN)} \end{pmatrix}$$

In the matrices $A$, $B$ and $A_B^i$, each row represents a set of parameter to be used as an input for the model. Numerical simulations are performed, and the output of the sample matrices $A$, $B$ and $A_B^i$ are stored as the vectors

$$Y_A = \begin{pmatrix} Y\left(A^{(A1)}\right) \\ Y\left(A^{(A2)}\right) \\ \vdots \\ Y\left(A^{(AN)}\right) \end{pmatrix}; \quad Y_B = \begin{pmatrix} Y\left(B^{(B1)}\right) \\ Y\left(B^{(B2)}\right) \\ \vdots \\ Y\left(B^{(BN)}\right) \end{pmatrix}; \quad Y_{A_B^i}. \tag{16}$$

where $Y_A$, $Y_B$ and $Y_{A_B^i}$ are output vectors.

The final step involves the calculation of the sensitivity indices, using the samples generated during the sampling scheme. We computed the total effect indices, given by

$$S_{T_i} = 1 - \frac{Y_A \cdot Y_B - f^2}{Y_A \cdot Y_A - f^2} \tag{17}$$

where $f$ is defined as

$$f := \frac{1}{N} \sum_{j=1}^{N} Y_A^{(j)}. \tag{18}$$

This index indicates the contribution of the parameter to the output of the model. The importance of each parameter $i$ is proportional to the value of $S_{T_i}$, meaning that higher $S_{T_i}$ leads to a higher contribution to the model output (*48*). Parameters with higher $S_T$ need a more carefully calibration, as small error during the calibration can lead to larger errors to the predictions generated by the model. The total effect takes into account higher-order interactions among model variables; thus, correlation between variables can also be identified using this method. In addition, we also evaluated the influence of first-order effects, which do not consider interactions among variables, to the model output.

The numerical simulations were performed using the SALib library (*48*). The experiment was conducted generating $N = 15{,}000$ parameter combinations, totaling $225{,}000$ simulations of the model, and the result shows the evolution of the parameters according to $S$, $E$, $I_a$, $I_s$, $U$, $H$, $R$ and $D$ compartments.

The results for the sensitivity analysis of the SEIIHURD+$\psi$ model confirm our previous findings of the sensitivity of the SEIIHURD model, where we showed that the transmission rate, $\beta$, and $\delta$, the factor that reduces the infectivity of asymptomatic/non-detected individuals, were among the most influential parameters to every model output during most of the evaluated periods (*9*). This result is recapitulated in every compartment throughout the course of the model dynamics studied herein (Fig. S7). In the initial simulation period (up to day 20), the parameters that exert the most impact to the compartments related to severe disease ($H$, $U$) and fatalities ($D$), after $\beta$ and $\delta$, was $h$ (the proportion of symptomatic needing hospitalization or ICU). In $U$, the parameter $\xi$ also appears to play an important role in the dynamics of critical cases, which is expected as the parameter $1 - \xi$ associates to the proportion of hospitalized symptomatic that proceed to ICU (Fig. S7E,F,H). The parameter $p$, the proportion of latent ($E$) that proceed to symptomatic infective, appears as an important factor governing the dynamics of individuals in $I_s$, specially at the initial simulation period and at later time points.

In line with our previous findings on the dynamics of the SEIIHURD model system (*9*), now expanded to the SEIIHURD+$\psi$ framework, the sensitivity analysis conducted confirmed the importance of carefully considering the intervals of $\beta$, $\delta$, $\xi$ and $h$, as these parameters represent important determinants of the dynamics of the model. Indeed, we have previously conducted an extensive literature mining of these key parameters to inform their ranges (table 2 and Ref. (*9*)).

#### NMPC Algorithm

---

**Finitely Parametrized NMPC for Social Distancing Guidelines**

---

**Initialize**: $N(0)$, $S(0)$, $E(0)$, $I_a(0)$, $I_s(0)$, $H(0)$, $U(0)$, $R(0)$ and $D(0)$.
**Require**: $Q$, $n_I$, $n_{\text{beds}}$, $n_{\text{ICU}}$
**Loop, every day**:

- Step (*i*): "Measure" the available contagion data ($I(k)$, $H(k)$, $U(k)$, $\psi(k)$ and $D(k)$);
- Step (*ii*): **Loop every week**:
  - Step (a): **For each control sequence** $j$:
    * Step (1): Build the control vector $\mathcal{U}_k$
    * Step (2): Explicitly simulate the future sequence of the SEIIHURD variables;
    * Step (3): Evaluate if constraints are respected. If they are not, end, else, compute the cost function $J_{\text{NMPC}}(\cdot)$ value.
  - **end**
  - Step (b): Choose the optimal control value $\mathcal{U}_k$ that corresponds to the smallest $J(\cdot)$.
  - Step (c): Increment $k$, i.e. $k \leftarrow k + 1$.
- **end**
- Step (*iii*): Apply the local control policy $u(k)$.
- Step (*iv*): Simulate the SEIIHURD+$\psi$ model.
- Step (*v*): Increment $k$, i.e. $k \leftarrow k + 1$.

**end**

---

### Supplementary Figures

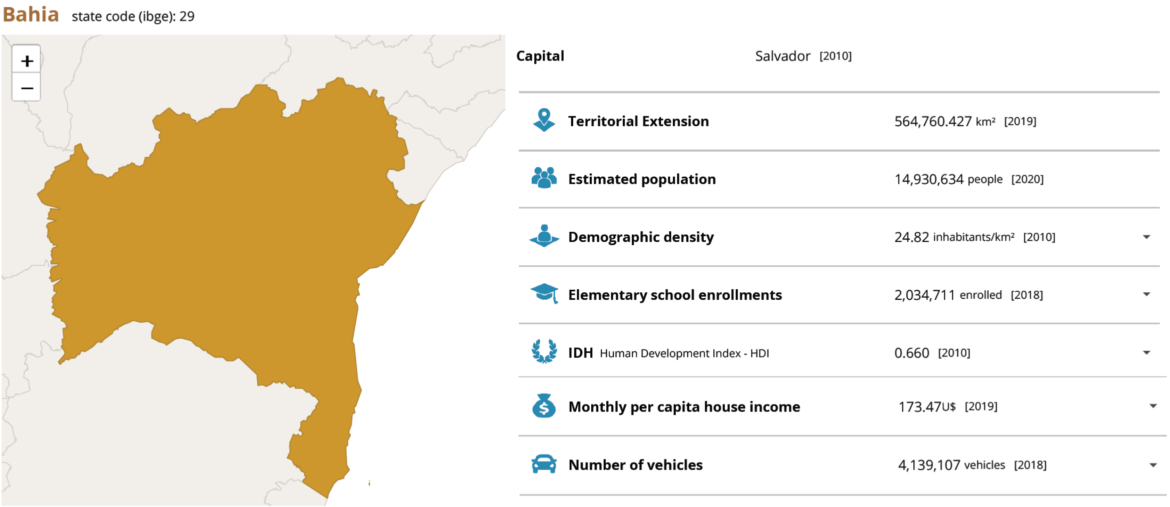

**Bahia** state code (ibge): 29

| | |
|---|---|
| Capital | Salvador [2010] |
| Territorial Extension | 564,760.427 km² [2019] |
| Estimated population | 14,930,634 people [2020] |
| Demographic density | 24.82 inhabitants/km² [2010] |
| Elementary school enrollments | 2,034,711 enrolled [2018] |
| IDH Human Development Index - HDI | 0.660 [2010] |
| Monthly per capita house income | 173.47U$ [2019] |
| Number of vehicles | 4,139,107 vehicles [2018] |

**Fig. S 1. A factsheet of Bahia, Brazil, with the main economic and social indicators**. Extracted from the Brazilian Institute of Geography and Statistics (IBGE). The exchange rate used to convert Brazilian reais (BRL) to US dollars (USD) was 1 BRL = 0.19 USD (as of Dec 29, 2020).

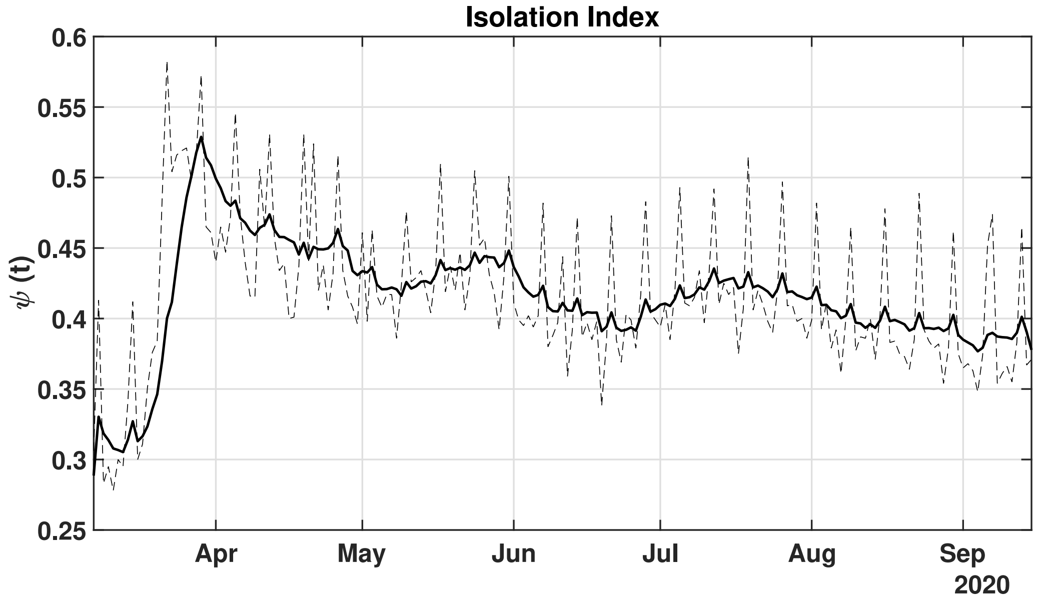

**Fig. S 2. Social mobility reduction index**. The dashed lines represent the daily percentage of social mobility given by InLoco. The full black line is the moving average mean of 8 days.

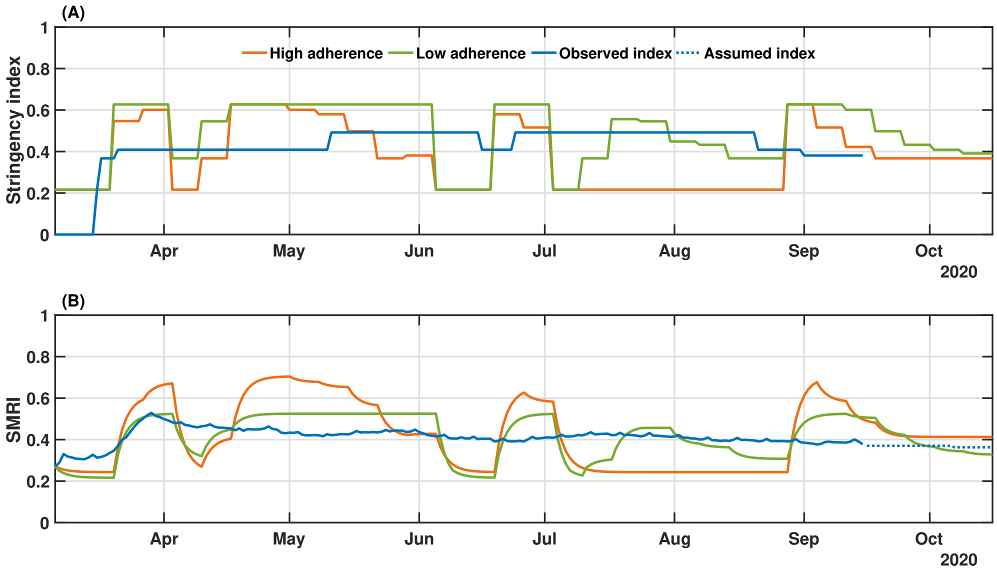

**Fig. S 3. Real and simulated social mobility and governmental interventions in the state of Bahia.** Levels of stringency (A) and social mobility reduction (B) indexes are shown for the high and low compliance scenarios, as well as the actual value of these metrics in the state-level during the period. The observed SMRI values (March 6-September 15) consist of a 8-day moving average. The dotted line in panel (B) indicates the assumed values of SMRI as described in Results. This scenario represents a hypothetical situation in which the government would have applied 21 decrees with stringency index varying between $0.216$ to $0.6269$, as described in Table S4.

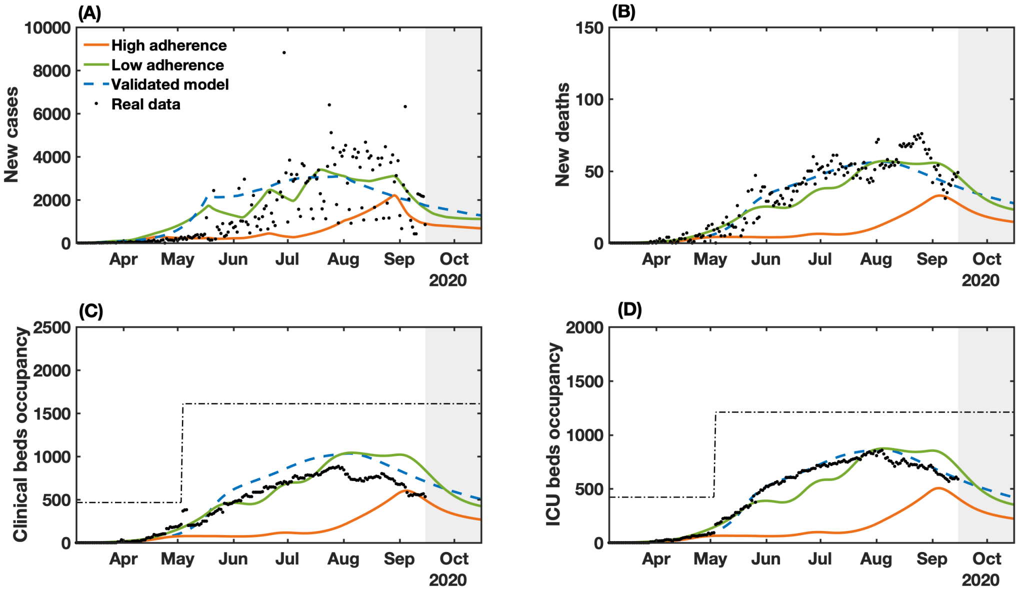

**Fig. S 4. Real and simulation-controlled COVID-19 epidemic unfolding in Bahia, Brazil.** (A) New cases; (B) deaths; (C) clinical hospitalization and (D) ICU bed requirements at the state level. The dashed-blue lines represent the dynamics of the validated model presented in Fig. 2 considering the observed SMRI time series in Fig. 3B. The dashed-dotted lines represent the clinical and ICU bed limits. Raw data (black dots) from March 6 to September 15, 2020 are shown in this graph. This scenario represent a hypothetical situation in which the government would have applied 21 decrees with stringency index varying between 21.62% and 62.69%, as described in Table S4.

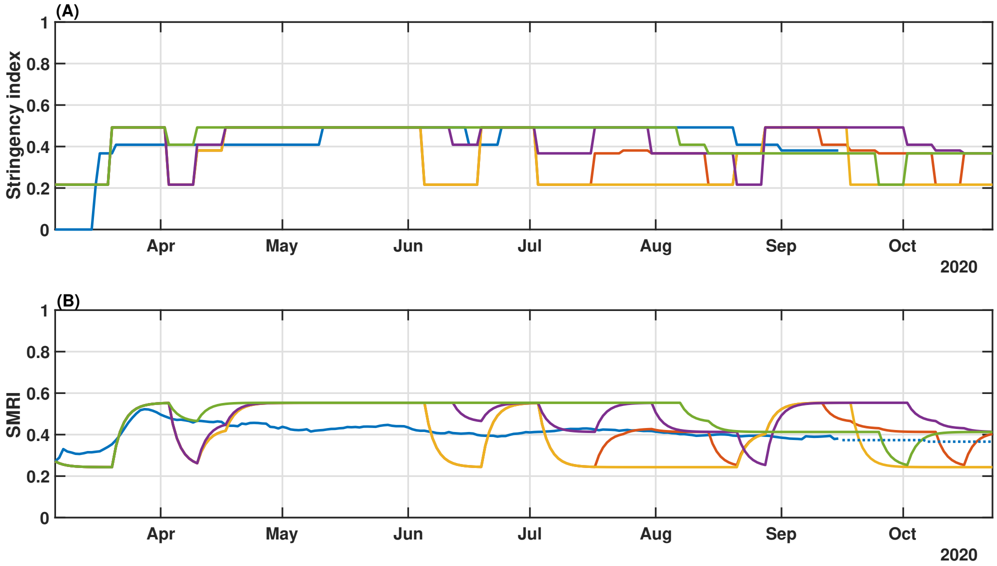

**Fig. S 5. Real and simulated social mobility and governmental interventions in the state of Bahia.** Levels of stringency (A) and social mobility reduction (B) indexes. The blue-solid line represents the real measured index applied in the state from March 16 to September 15. The dotted line in panel (B) indicates the assumed values of SMRI. This scenario simulates different control tuning with the limited variation of the stringency index between 21.62% and 49.21%, changing the parameter $Q$ to adjust the trade-off between reducing the level of the measures or minimizing the number of cases. $Q$ was adjusted according to the time windows: i) from March 6 to May 15 ii) from May 16 to August 23 and iii) from August 24 to October 15. For the red line $Q$ is $8e4, 3e4, 1e4$. For the yellow line $Q$ is $5e6, 1e6, 2e5$. For the violet line $Q$ is $1e4, 5e3, 1e3$. For the green line $Q$ is $5e3, 1e3, 1e3$.

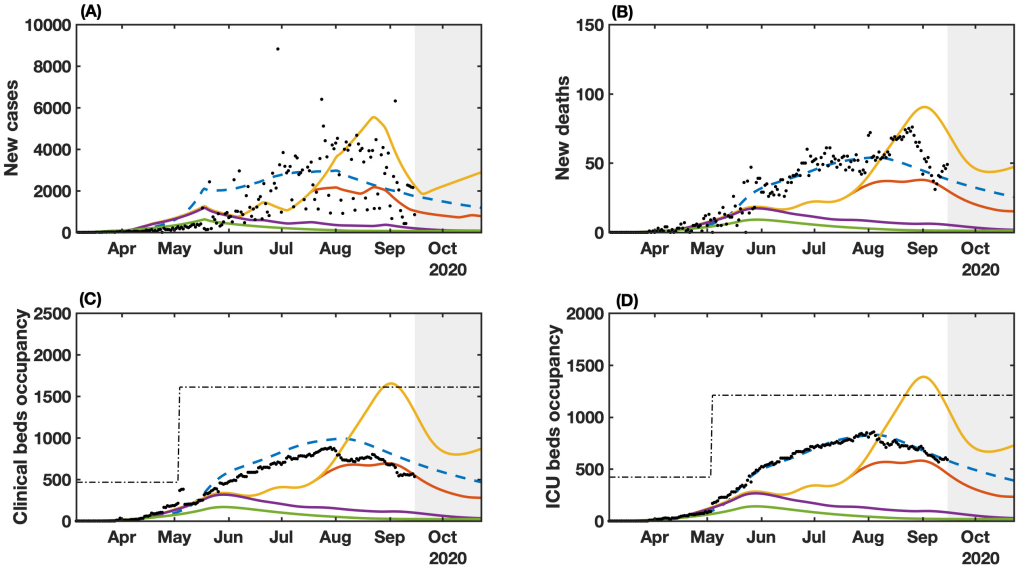

**Fig. S 6. Real and simulation-controlled COVID-19 epidemic behavior in Bahia, Brazil.** (A) New cases; (B) deaths; (C) clinical hospitalization and (D) ICU bed requirements at the state level. The dashed-blue lines represent the dynamics of the validated model presented in Fig. 2 considering the observed SMRI time series in Fig. 3B. The dashed-dotted lines represent the clinical and ICU bed limits. Raw data (black dots) from March 6 to September 15, 2020 are shown in this graph. This scenario simulates different control tuning with the limited variation of the stringency index between 21.62% and 49.21%, changing the parameter $Q$ to adjust the trade-off between reduce the level of the measures or minimize the number of cases. The $Q$ was adjusted accordingly to the time windows: i) from March 6 to May 15 ii) from May 16 to August 23 and iii) from August 24 to October 15. For the red line $Q$ is $8e4, 3e4, 1e4$. For the yellow line $Q$ is $5e6, 1e6, 2e5$. For the violet line $Q$ is $1e4, 5e3, 1e3$. For the green line $Q$ is $5e3, 1e3, 1e3$.

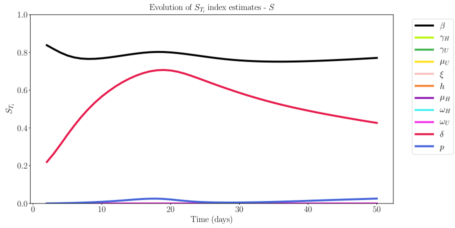

(A) $S_{T_i}$ for $S$

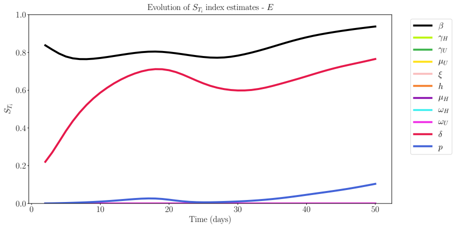

(B) $S_{T_i}$ for $E$

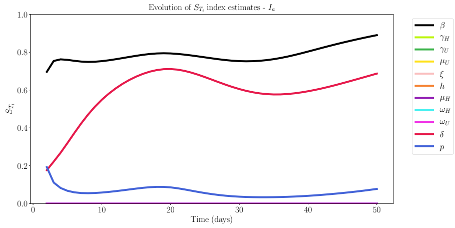

(C) $S_{T_i}$ for $I_a$

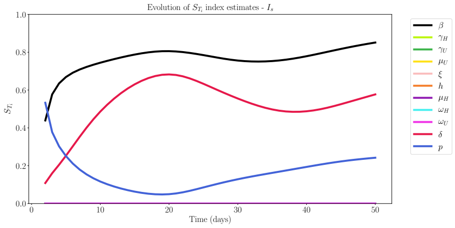

(D) $S_{T_i}$ for $I_s$

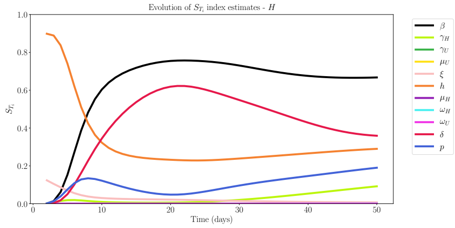

(E) $S_{T_i}$ for $H$

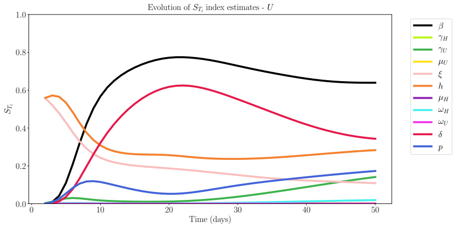

(F) $S_{T_i}$ for $U$

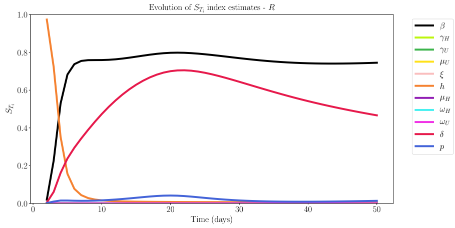

(G) $S_{T_i}$ for $R$

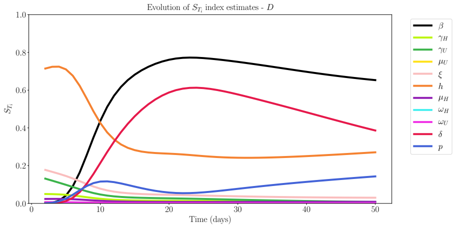

(H) $S_{T_i}$ for $D$

**Fig. S 7.** Sensitivity analysis study for $S$, $E$, $I_a$, $I_s$, $U$, $H$, $R$ and $D$ compartments over time. The total effect index $S_{T_i}$ is shown for each evaluated parameter in each compartment of the SEIIHURD model.

(A)

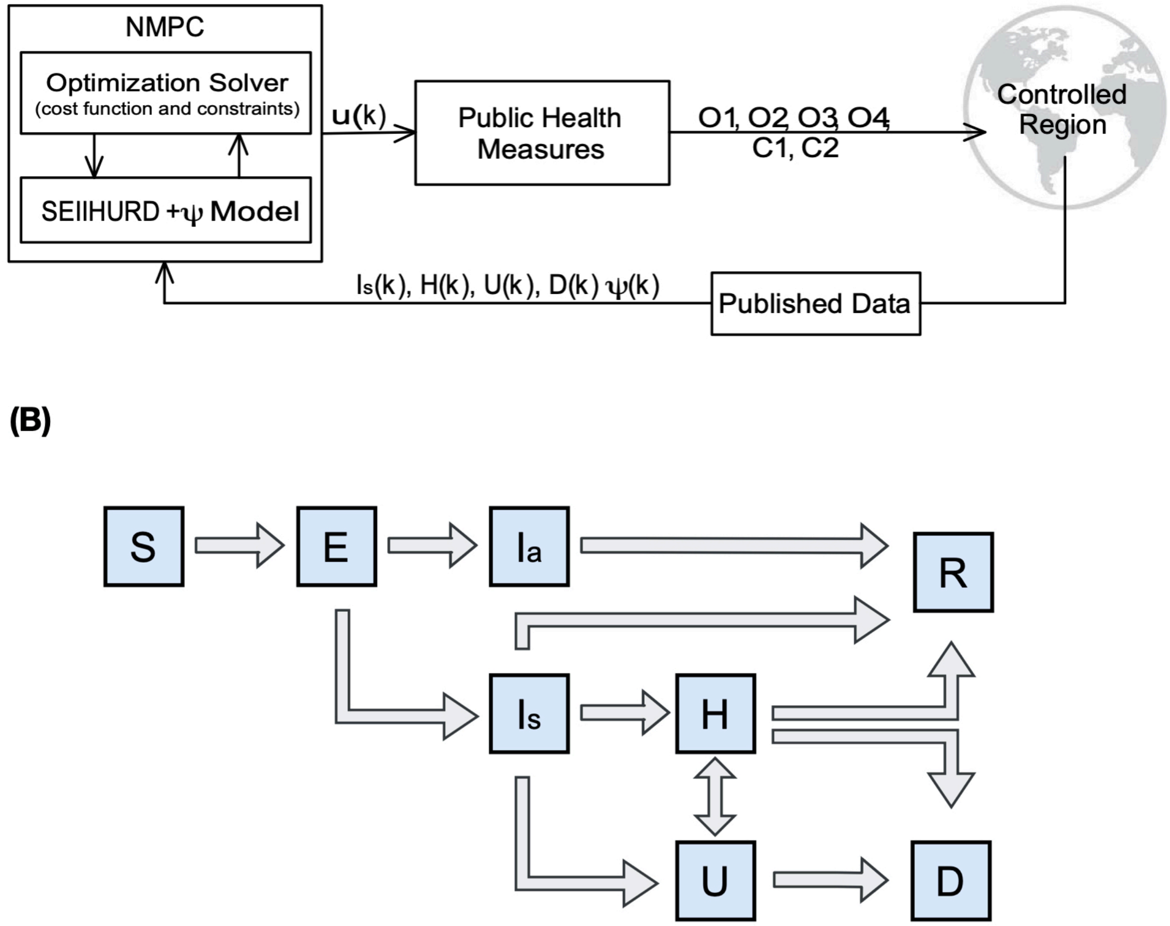

**Fig. S 8. (A) Control loop scheme and (B) SEIIHURD compartmental model.** The notation $k$ used in (A) defines the discrete sample time in the control algorithm.

### Supplementary Tables

**Table S 1. Categories of governmental measures to mitigate the spread of COVID-19 in Bahia.** Details are described in the work of Jorge et al. (*7*).

| Measure ID | Guideline | Interval |
|---|---|---|
| O1 | Cancel public events | [0,1] |
| O2 | Closure of schools/universities | [0,1] |
| O3 | Home-office labor for government employees | [0,1] |
| O4 | Social Isolation | [0,1] |
| C1 | Closure of non-essential activities (business, cultural activities, etc) | [0,1] |
| C2 | Transport lockdown | [0,1] |

**Table S 2. Key epidemiological parameters used in the SEIIHURD+$\psi$ model without gains**. Estimates and values for the analysis of the SEIIHURD+$\psi$ without gains with their search interval (when applicable) and respective estimates obtained according to best fit to data up to September 15, 2020 for Bahia. The parameters search intervals were informed by our previous literature mining (*9*).

| Parameter | Description | Interval | Fixed | Estimated (95% CI) |
|---|---|---|---|---|
| $\beta_0$ | Pre-intervention transmission rate | $[0, 3]$ | - | 2.13 (2.02-2.24) |
| $\beta_1$ | Post-intervention transmission rate | $[0, 3]$ | - | 1.76 (1.67-1.85) |
| $\beta_2$ | Post-intervention transmission rate | $[0, 3]$ | - | 1.13 (1.07-1.19) |
| $\beta_3$ | Post-intervention transmission rate | $[0, 3]$ | - | 1.00 (0.95-1.05) |
| $t_1$ | Time of transmission rate change | [March 26$^{th}$, May 17$^{th}$] | - | March 26 |
| $t_2$ | Time of transmission rate change | [May 18$^{th}$, August 1$^{st}$] | - | May 18 |
| $t_3$ | Time of transmission rate change | [August 1$^{st}$, September 15$^{th}$] | - | Aug 1 |
| $\delta$ | Asymptomatic/non-detected infectivity factor | [0,0.75] | 0.31 | - |
| $p$ | Proportion of latent (E) that proceed to symptomatic infective | $[0.13, 0.5]$ | 0.2 | - |
| $\kappa$ | Mean exposed period | $[1/6, 1/3]$ | $1/4$ | - |
| $\gamma_a$ | Mean asymptomatic period | $[1/3.70, 1/3.24]$ | $1/3.5$ | - |
| $\gamma_s$ | Mean symptomatic period | $[1/5, 1/3]$ | $1/4$ | - |
| $h$ | Proportion of symptomatic needing hospitalization or ICU | $[0.05, 0.25]$ | 0.06 | - |
| $1-\xi$ | Proportion of symptomatic that proceed to ICU | $[0.01, 0.5]$ | 0.47 | - |
| $\gamma_H$ | Mean hospitalization (clinical beds) period | $[1/12, 1/4]$ | 0.18 | - |
| $\gamma_U$ | Mean ICU period | $[1/12, 1/3]$ | 0.13 | - |
| $\mu_H$ | Death rate of hospitalized individuals | $[0.1, 0.2]$ | 0.15 | - |
| $\mu_U$ | Death rate of ICU individuals | $[0.4, 0.5]$ | 0.4 | - |
| $\omega_H$ | Proportion of hospitalized that goes to ICU | $[0.1, 0.3]$ | 0.14 | - |
| $\omega_U$ | Proportion of ICU that goes to hospitalization | $[0.1, 0.3]$ | 0.29 | - |

**Table S 3. Parameter specifications for the optimizations procedure.**

| Parameter | Search interval | Initial condition for the optimization problem |
|---|---|---|
| $g_1\beta$ | [0; 3] | 1 |
| $g_2 h$ | [0.05; 0.25] | 0.06 |
| $g_3\xi$ | [0.5; 0.99] | 0.53 |
| $g_4\mu_U$ | [0.4; 0.5] | 0.4 |
| $g_5\gamma_U$ | [1/12; 1/3] | 0.13 |
| $g_6\gamma_H$ | [1/12; 1/4] | 0.18 |
| $g_7 p$ | [0.13; 0.5] | 0.2 |
| $g_8\omega_U$ | [0.1; 0.3] | 0.29 |
| $g_9\omega_H$ | [0.1; 0.3] | 0.14 |
| $g_{10}\mu_H$ | [0.1; 0.2] | 0.15 |
| $g_{11}\delta$ | [0.01; 0.75] | 0.31 |

**Table S 4. Control Input evaluation**

| Control Input Value | Social Distancing Measure ID |
|---|---|
| $0\,\%$ | Do nothing |
| $21.62\,\%$ | O1 = $0.333$, O2 = $0.5$, O3 = $0.25$, O4 = $0.0$, C1 = $0.214$ and C2 = $0.00$ |
| $36.71\,\%$ | O1 = $0.667$, O2 = $1.0$, O3 = $0.25$, O4 = $0.0$, C1 = $0.286$ and C2 = $0.00$ |
| $38.10\,\%$ | O1 = $0.667$, O2 = $1.0$, O3 = $0.25$, O4 = $0.0$, C1 = $0.428$ and C2 = $0.00$ |
| $39.00\,\%$ | O1 = $0.500$, O2 = $1.0$, O3 = $0.25$, O4 = $0.0$, C1 = $0.286$ and C2 = $0.25$ |
| $40.87\,\%$ | O1 = $0.667$, O2 = $1.0$, O3 = $0.25$, O4 = $0.0$, C1 = $0.286$ and C2 = $0.25$ |
| $41.50\,\%$ | O1 = $0.667$, O2 = $1.0$, O3 = $0.39$, O4 = $0.0$, C1 = $0.428$ and C2 = $0.00$ |
| $42.23\,\%$ | O1 = $0.667$, O2 = $1.0$, O3 = $0.39$, O4 = $0.0$, C1 = $0.456$ and C2 = $0.01$ |
| $43.25\,\%$ | O1 = $0.667$, O2 = $1.0$, O3 = $0.25$, O4 = $0.0$, C1 = $0.428$ and C2 = $0.25$ |
| $44.84\,\%$ | O1 = $0.333$, O2 = $1.0$, O3 = $1.00$, O4 = $0.0$, C1 = $0.357$ and C2 = $0.00$ |
| $45.90\,\%$ | O1 = $0.667$, O2 = $1.0$, O3 = $0.39$, O4 = $0.0$, C1 = $0.442$ and C2 = $0.25$ |
| $47.05\,\%$ | O1 = $0.667$, O2 = $1.0$, O3 = $0.39$, O4 = $0.0$, C1 = $0.511$ and C2 = $0.25$ |
| $49.21\,\%$ | O1 = $0.667$, O2 = $1.0$, O3 = $0.25$, O4 = $0.5$, C1 = $0.286$ and C2 = $0.25$ |
| $49.80\,\%$ | O1 = $0.333$, O2 = $1.0$, O3 = $1.00$, O4 = $0.0$, C1 = $0.571$ and C2 = $0.08$ |
| $51.59\,\%$ | O1 = $0.667$, O2 = $1.0$, O3 = $0.25$, O4 = $0.5$, C1 = $0.428$ and C2 = $0.25$ |
| $54.56\,\%$ | O1 = $0.333$, O2 = $1.0$, O3 = $1.00$, O4 = $0.0$, C1 = $0.857$ and C2 = $0.08$ |
| $54.69\,\%$ | O1 = $0.667$, O2 = $1.0$, O3 = $0.39$, O4 = $0.5$, C1 = $0.469$ and C2 = $0.25$ |
| $55.61\,\%$ | O1 = $0.667$, O2 = $1.0$, O3 = $0.39$, O4 = $0.5$, C1 = $0.525$ and C2 = $0.25$ |
| $57.94\,\%$ | O1 = $0.667$, O2 = $1.0$, O3 = $1.00$, O4 = $0.0$, C1 = $0.642$ and C2 = $0.16$ |
| $60.12\,\%$ | O1 = $0.667$, O2 = $1.0$, O3 = $1.00$, O4 = $0.0$, C1 = $0.857$ and C2 = $0.08$ |
| $62.69\,\%$ | O1 = $0.667$, O2 = $1.0$, O3 = $1.00$, O4 = $0.0$, C1 = $0.928$ and C2 = $0.16$ |

**Table S 6. Key epidemiological parameters used in the SEIIHURD model.**

| Parameter | Description |
|---|---|
| $\beta$ | Transmission rate |
| $t_i$ | Time of transmission rate change implicit on the Heaviside step function of $\beta$ |
| $\delta$ | Asymptomatic/non-detected infectivity factor |
| $p$ | Proportion of latent (E) that proceed to symptomatic infective |
| $\kappa$ | Mean exposed period (days$^{-1}$) |
| $\gamma_a$ | Mean asymptomatic period (days$^{-1}$) |
| $\gamma_s$ | Mean symptomatic period (days$^{-1}$) |
| $h$ | Proportion of symptomatic needing hospitalization (clinical beds) or ICU |
| $1 - \xi$ | Proportion of symptomatic that proceed to ICU |
| $\gamma_H$ | Mean hospitalization (clinical beds) period (days$^{-1}$) |
| $\gamma_U$ | Mean ICU period (days$^{-1}$) |
| $\mu_H$ | Death rate of hospitalized individuals |
| $\mu_U$ | Death rate of ICU individuals |
| $\omega_H$ | Proportion of hospitalized that goes to ICU |
| $\omega_U$ | Proportion of ICU that goes to hospitalization |

**Table S 5. Identified epidemiological parameters used in the SEIIHURD+$\psi$ model with gains**. Identified parameter values coupled with its respective gain using the optimization method for a time window of 13 days, starting from March 6, 2020

| | Time Window | | | | | | | | | | | | |
|---|---|---|---|---|---|---|---|---|---|---|---|---|---|
| Gains | 1 | 2 | 3 | 4 | 5 | 6 | 7 | 8 | 9 | 10 | 11 | 12 | 13 |
| $g_1\beta$ | 0.5486 | 1.7002 | 3.0000 | 2.2302 | 0.6286 | 0.9703 | 1.1252 | 1.2230 | 1.2148 | 0.6290 | 0.7149 | 2.5589 | 0.5730 |
| $g_2h$ | 0.0500 | 0.2500 | 0.2221 | 0.2500 | 0.2500 | 0.1947 | 0.1303 | 0.0568 | 0.0627 | 0.0500 | 0.0847 | 0.0500 | 0.0500 |
| $g_3\xi$ | 0.5000 | 0.6319 | 0.5000 | 0.7181 | 0.6143 | 0.8228 | 0.5000 | 0.5000 | 0.5000 | 0.5107 | 0.6052 | 0.5000 | 0.5000 |
| $g_4\mu_U$ | 0.5000 | 0.4000 | 0.4000 | 0.4000 | 0.5000 | 0.5000 | 0.5000 | 0.5000 | 0.5000 | 0.5000 | 0.4000 | 0.4984 | 0.5000 |
| $g_5\gamma_U$ | 0.3333 | 0.0833 | 0.1814 | 0.0932 | 0.0833 | 0.0833 | 0.1001 | 0.0875 | 0.0972 | 0.1631 | 0.1328 | 0.1232 | 0.1521 |
| $g_6\gamma_H$ | 0.2500 | 0.0833 | 0.2500 | 0.0833 | 0.1018 | 0.2500 | 0.1379 | 0.1028 | 0.1169 | 0.1902 | 0.2500 | 0.1923 | 0.1905 |
| $g_7p$ | 0.1300 | 0.2548 | 0.1750 | 0.2732 | 0.4033 | 0.5000 | 0.1969 | 0.2671 | 0.2062 | 0.1300 | 0.1300 | 0.1626 | 0.1796 |
| $g_8\omega_U$ | 0.1000 | 0.3000 | 0.3000 | 0.3000 | 0.1000 | 0.1000 | 0.1000 | 0.1000 | 0.1000 | 0.1000 | 0.3000 | 0.2004 | 0.1000 |
| $g_9\omega_H$ | 0.1000 | 0.3000 | 0.3000 | 0.3000 | 0.3000 | 0.3000 | 0.3000 | 0.3000 | 0.3000 | 0.3000 | 0.1000 | 0.1000 | 0.1891 |
| $g_{10}\mu_H$ | 0.1000 | 0.1000 | 0.2000 | 0.2000 | 0.2000 | 0.1020 | 0.2000 | 0.2000 | 0.2000 | 0.1000 | 0.1000 | 0.1000 | 0.1000 |
| $g_{11}\delta$ | 0.7500 | 0.7500 | 0.0779 | 0.0100 | 0.7500 | 0.7500 | 0.7500 | 0.0355 | 0.7500 | 0.7500 | 0.7500 | 0.0100 | 0.7500 |
